# Interactions between Perceived Stress and Microbial-Host Immune Components in Pregnancy

**DOI:** 10.1101/2022.05.08.22274745

**Authors:** Beatriz Peñalver Bernabé, Pauline M. Maki, Janet L. Cunningham, Tory Eisenlohr-Moul, Lisa Tussing-Humphreys, Ian M. Carroll, Samantha Meltzer-Brody, Jack A. Gilbert, Mary Kimmel

## Abstract

**Background:** Higher stress during pregnancy associates with negative outcomes and elevated inflammation. The gut microbiota reflects the environment, lifestyle, and the perinatal period. The gut microbiota, alongside host immune responses, has the potential to aid in identifying when stress is excessive.

**Methods:** Two U.S. cohorts, of 84 pregnant individuals, composed of urban women of color and suburban white women, completed the Perceived Stress Scale-10 (PSS-10) and provided fecal and blood samples at two time points. Confirmatory Factor Analysis assessed the robustness of a two-factor PSS-10 (Emotional Distress and Self-Efficacy) in the two cohorts. Gut microbiota composition was measured by 16S rRNA amplicon sequencing and the immune system activity was assessed with a panel of 21 T-cell related cytokines and chemokines.

**Results:** Emotional Distress levels were higher in the suburban compared to the urban cohort, but levels of Self-Efficacy were similar. Emotional Distress and Self-Efficacy levels were associated with distinct taxonomical signatures and the gut microbiota data improved the prediction of Self-Efficacy levels compared with models based on socio-demographic characteristics alone. Integration of self-reported symptoms, microbial and immune information revealed a possible mediation effect of *Bacteroides uniformis* between the immune system (through CXCL11) and Self-Efficacy.

**Conclusions:** The study identified links between distinct taxonomical and immunological signatures with perceived stress. The data is congruent with a model where gut microbiome and immune factors may modulate neural plasticity resulting in increased Self-Efficacy during pregnancy. The predictive value of these peripheral markers merit further study.

## INTRODUCTION

Pregnancy is a time of diverse biological, social and psychological adaptation.^1,2^ High perceived stress, defined by Cohen as nervousness, fear, and anger resulting from perceiving that one’s life is out of control with troubles that one can’t overcome,^3^ during pregnancy increases the risk for development of perinatal neuropsychiatric symptoms, including anxiety, depressed mood, psychosis, and substance use disorders.^4–6^ High perceived stress during pregnancy has also been associated with obstetrical complications, such as gestational diabetes mellitus, preterm birth, gestational hypertension, preeclampsia and complications for the infant, such as being small for gestational age.^7–11^

The way that stress is experienced and expressed and the resulting physical and mental health effects is highly individualized, and may be influenced by culture. The data-driven Woods-Giscombe’s Superwoman schema explains that Black women feel an obligation to manifest strength and independence, suppress emotions, including reporting perceived stress, succeed despite limited resources, and help others,^12^ but the combined effects of structural inequities and chronic experienced stress explain poorer health.^12^ For example, Black women and Latinas are at much higher risk of obstetrical complications (e.g., preterm birth, gestational diabetes mellitus, pre-eclampsia),^13^ but they report lower levels of overall perceived stress.^14^ In a study of U.S. non-pregnant individuals with over 56% female who self-identifying as Mexican-origin, participants that reported higher perceived stress had lower 10-year cardiovascular disease risk, suggesting that in some populations expressing stress may be more acceptable and result in improved health outcomes.^15^ This indicates that self-reported perceived stress is a complex construct that could be impacted by cultural and environmental factors. Thus these factors should be carefully considered when using self-reported measurements of perceived stress to asses risk of negative outcomes in distinct populations.

Biological markers that accurately reflect stress levels that have negative health effects could serve to identify those individuals at risk for perinatal complications.^16–18^ There is increasing evidence that the microbiota-gut-brain axis, that encompasses interactions between the neurological, endocrinological and immunological systems, is associated with mental health disorders and obstetrical complications.^10,19–24^ The microbiota that reside in the gastrointestinal system can be influenced by lifestyle (e.g., diet, exercise), residential location and home environment (e.g., rural versus urban), and medication use (e.g., antibiotics).^25^ Disruptions in the gut microbial ecosystem have been associated with both intestinal and systemic diseases (e.g., IBS, diabetes, depression).^26^ Initial cross sectional studies in pregnancy indicate an association between gut microbial communities, maternal psychosocial stress, and host immune system responses.^27,28^ Osborne, et. al. found an increase in pro-inflammatory innate immunity cytokines in the third trimester in individuals with higher depression and anxiety.^29^ Important to the offspring’s development, maternal stress during pregnancy influences the infants’ gut microbiome composition.^23,30^ The integration of gut microbiome and markers of the immune system, such as circulating cytokines and chemokines, could be potential ideal biological biomarkers of perceived stress and thus of negative perinatal outcomes.

We recruited a total of 84 pregnant individuals from two sociodemographic and geographically distinct U.S. locations, women of color from an urban population in the Midwest and White women from a suburban population from the mid-Atlantic region. We aimed to understand the maternal gut-microbiota-brain axis during the second and third trimesters of pregnancy through the integration of maternal gut microbiota composition, maternal cytokine and chemokine concentrations in serum, and self-reported perceived levels of stress in these two distinct cohorts. We hypothesized that models that included maternal gut microbiota and systemic inflammatory characteristics will better predict perceived stress levels during pregnancy than those models based on socio-demographic characteristics alone.

## MATERIALS AND METHODS

### Participant Recruitment

Our sample included US pregnant individuals from two sociodemographic and geographically distinct cohorts: one a sample from a Midwest large city, Chicago (Illinois), comprising urban-dwelling women of color (Black women and Latinas) with low socioeconomic status (SES) and low education, and the other a sample from mid-Atlantic city, Chapel Hill (North Carolina), comprising suburban White women with a higher SES and level of education. Urban participants were recruited during their initial obstetric visit at a public university hospital before 16 weeks gestational age (urban cohort). Suburban participants were recruited through advertisement and their initial visit was before 28 weeks gestational age. Participants provided fecal samples during the second trimester (T2, range=24-29 weeks gestational age); and the third trimester (T3, range=33-38 weeks gestational age) and blood samples at the T2 visit. Study protocols were approved by the University of Illinois Chicago, Chicago, IL, US (IRB# 2014-0325, IRB# 2018-0842) and the University of North Carolina, Chapel Hill, NC, US (IRB# 16-0959, IRB# 16-2783) Institutional Review Boards.

### Self-reported mental health questionnaires

At each visit, participants completed questionnaires about demographics and the self-assessment tools including the Generalized Anxiety Disorder-7 (GAD-7),^31^ Perceived Stress Scale (PSS)-10,^3,32^ and either the Patient Health Questionnaire (PHQ-9)^33)^ in the urban (Chicago) or the Edinburgh Postnatal Depression Scale (EPDS)^34^ in the suburban (Chapel Hill) cohorts. Participants also consented to access their electronic medical records. The PSS-10 is one of the most common self-report tools used to assess perceived stress.^3,32^ The PSS-10 is a 10-item self-reported questionnaire.^3^ Each question is scored on a 5-point Likert scale ranging from 0 “*Never*” to 4 “*Very often*. “

### Confirmatory Factor Analysis (CFA)

In both non-pregnant and pregnant individuals, total PSS-10 scores have been consistently divided into two factors, “perceived helplessness” or “perceived distress” and “perceived self-efficacy”^35^ or “perceived coping”.^36,37^ Perceived distress and perceived self-efficacy are inversely correlated, providing information about the balance between perceived distress and perceived resilience. Here, we also estimated the ratio between Emotional Distress and Self-Efficacy (Ratio). To compare the two PSS-10 subconstructs, Emotional Distress and Self-Efficacy, in the two cohorts, we performed CFA using latent variable modeling implemented in the R package *lavaan*.^38^ We explored the fitting of the two-latent variable perceived stress model by model configural, metric (loading), scalar (intercept) and residual invariances. Models with comparative fit index (CFI) and Tucker-Lewis Index (TLI) greater than 0.95 and root mean square error of approximation (rmsea) less than 0.05 were deemed appropriate for fitting the data. Nested models were compared with ANOVA using a statistical significant p-value cutoff of less than 0.05. (See Supplemental methods for more details).

### Reduced PSS-10

To identify the core PSS-10 items that support model structural invariance between the two cohort sites (urban and suburban), we employed an iterative process in which one item was removed at a time until the model metric, scalar and residual invariabilities were maintained between the sites. We employed the score test or Lagrange Multiplier test, which releases one fixed or constrained parameter in the model at a time. The PSS-10 item with the lowest p-value was removed from the model until the ANOVA comparisons between the nested models was no longer statistically significant (p>0.05). Analyses were performed in R using the “*lavaan*” package.^38^

### Exploratory factor analysis

Correlations between the Likert scores for each PSS-10 item were calculated using Polychoric correlation with an oblique rotation, “Promax”, as PSS-10 Likert scores are highly correlated. The number of dimensions was selected using paralleled analysis with maximum likelihood as factoring method. Loadings for each question were estimated using minimum residuals with 2,000 iterations. Internal consistency between the self-reported questions assigned in each symptom dimension was determined with Cronbach’s alpha.^39^ Analyses were performed in R using the “*psych*” package.^40^

### Cytokine analysis

Blood samples were collected by a trained phlebotomist during a clinical or research visit, processed for serum, then aliquoted and stored at -80 °C. Samples were analyzed in duplicate at the University of Chicago (Chicago, IL, US) using a T-cell specific panel (HSTCMAG28SPMX21 from Milliplex®) that simultaneously analyzes CX3CL1 (Fractalkine), GM-CSF, IFNγ, IL-1β, IL-2, IL-4, IL-5, IL-6, IL-7, IL-8, IL-10, IL-12 (p70), IL-13, IL-17A, IL-21, IL-23, CXCL11 (ITAC), CCL3 (MIP-1α), CCL4 (MIP-1β), CCL20 (MIP-3α), TNF-α.

### Fecal sample collection

Most of the collected fecal samples from the urban cohort were rectal swabs (96%) and just 3 samples, each for 3 different participants, were provided as stool. Samples were processed and stored at -80°C within two hours of collection. Stool samples from the suburban cohort were collected by participants at home, put on ice and transported within 24 to 48 hours to the laboratory. Stool specimens from both locations were homogenized, aliquoted, and then frozen at -80°C.

### Fecal DNA extraction and sequencing

DNA was extracted with QIAGEN® MagAttract® PowerSoil® DNA KF Kit and followed a slightly-modified manufacturing protocol,^41^ as previously described.^42^ The V4 region of the 16S rRNA gene of the barcoded fecal samples was targeted with region-specific primers, 515F-806R. Samples were sequenced on a 151bp x 12bp x 151bp MiSeq run. Empty blanks were included to control for external and cross-contamination: i) clean rectal swabs to identify possible contamination in the swabs employed in the urban population, n=16; ii) open rectal swabs exposed to the biosafety cabinet environment during the extraction and barcoding protocol, n=16; iii) empty vials to identify possible contamination in the extraction kits and downstream processing, n=35. All the samples were extracted and barcoded at the University of Chicago (Illinois, US).

### Sequence identification and filtering

Amplicon Sequence Variants (ASV) were determined with DADA2, using default parameters unless indicated otherwise.^43^ Paired reverse and forward sequences were filtered and truncated to a maximum of 150 base-pair length with maximum expectation of 0.75. Out of the 186 samples, ten samples were removed due to low number of reads (<5 reads). Of the removed samples, five were controls, two and three samples belonged to the urban and suburban controls, respectively. After inference and chimeric removal, only sequences whose lengths were between 251 to 254 bp were used for downstream analysis. Taxonomy was assigned using the Silva database v.132 (ref^44^). ASVs coming from contamination were removed. We assumed that ASVs that were not present in at least 90% of the samples (excluding controls) were contaminants. Samples whose relative abundance was less than 1% and all their reads were below 10 counts were also removed. All analyses were conducted in R.^45^

### Cytokine and chemokines analysis

Correlations between T-cell cytokines and chemokines concentrations in serum, in logarithmic scale, were estimated using Spearman partial correlations corrected by participant’s BMI and level of education (as a proxy for recruitment site). BMI was obtained either by participant’s electronical medical records (urban) or by researchers (suburban). Participants’ BMI was regressed out from the T-cell cytokines and chemokines log-normalized concentrations. Clustering was performed with k-means. Correlations between T-cell cytokines and chemokines concentrations, in logarithmic scale, with perceived stress dimensions (Emotional Distress, Self-Efficacy and Perceived Stress) was determined with Spearman correlations and also using linear models corrected for participant’s BMI.

### Gut microbiome analysis

Counts were normalized using cumulative sum scaling normalization (CSS).^46^ Alpha-and beta-diversity were calculated with the R package *phyloseq*.^47^ For alpha-diversity, we employed total observed ASV and Chao to estimated richness and Shannon, Inverse Simpson and Abundance-based Coverage Estimator (ACE) to estimate evenness. Volatility was determined as the change in beta-diversity, as calculated by Bray-Curtis and by normalized UNIFRAC distances, between the second and third trimester visit. Distances calculated with UNIFRAC^48^ were not based on CSS-normalized counts but rather by sample rarefication at 6,000 counts. Statistically significant differences in alpha-and beta-diversity with respect to perceived stress dimensions (Emotional Distress, Self-Efficacy and Perceived Stress) were determined with PERMANOVA, adjusting by research visit and recruitment site. Volatility differences as a function of gestational weeks differences between the second trimester and third trimester visits were assed with linear models correcting by recruitment site. Associations between perceived stress dimensions (Emotional Distress, Self-Efficacy, Perceived Stress and Perceived Stress Ratio) and ASVs were calculated using zero-inflated Generalized Linear Models adjusted by recruitment site and participant’s age and the

log2 CSS normalization factors for each sample using the R package *metagenomeSeq*.^49^ We employed false discovery rate to account for multiple comparisons. For T-related cytokines and chemokines abundance in serum, models were also corrected by participants’ BMI, age and recruitment site. Unless stated otherwise, all p-values were corrected by multiple comparisons using false discovery rate.

### Linear Mixed Models (LMM)

Following Rothschild and colleagues, we employed LMM to identify the predictability power of the gut microbiome data (Microbiome Associated Index) of the different reduced dimensions identified with factor analysis.^50^ Normalized ASV counts were summarized at the genus levels to construct the taxa kinship matrix (*KM)*. LMM for reduced Emotional Distress scores, PSS-10 scores and Ratio between Emotional Distress and Self-Efficacy were corrected by race, while for reduced Self-Efficacy scores were corrected by nulliparity. We developed LMMs for each of the dimensions, with and without the microbiota kinship matrix, using participants as random variables. Participants that didn’t have one measurement per visit were excluded. Models were validated using a 10 times cross-validation approach (see Supplemental Methods for more details).

### Co-abundances networks

ASVs were summarized at the genus levels. Correlations between CSS normalized genera were obtained with SparcCC.^51^ Edge significance was determined with 100 bootstrapping iterations using as a null-model a matrix created by scrambling the count data. Edges were deemed significant if their correlation coefficient and their z-scores were at least 3 times greater than the standard deviations of the corresponding bootstrapping correlations and z-scores for the sample edge.

### Statistical analysis

Unless otherwise specified, chi-square tests were used for comparisons of categorical variables, t-tests for categorial and continuous variables, and Spearman partial and non-particial correlations for continuous variables. All the p-values were corrected for multiple comparisons using false discovery rate (fdr).^52^ Analysis was conducted in R (ref^45^) and figures were produced with *ggplot2*.^53^

## RESULTS

### Comparison of socio-demographic characteristics

The urban cohort comprised more than 60% Black women and approximately 31% Hispanic women or Latinas. The suburban cohort comprised 78% non-Hispanic White women. Among the suburban cohort, 61% reported an education status of college or above, 96% were married or in committed relationship, and 28% were obese. This compared to women in the urban cohort in which only 16% completed some college education, 60% were married or in a committed relationship, and more than 44% were obese (**Table 1**).

**Table 1.** Demographic characteristics of the urban and suburban cohorts. N, number of participants; BMI, body mass index; false discovery rate corrected p values. Gestational weeks at the T2 [24, 29] and at the T3 [33,38] visits. ^**^fdr-corrected p <0.001; ^*^fdr-corrected p <0.01

|  | **Urban**<br>**N=38** | **Suburban**<br>**N=46** | **Urban**<br>**%** | **Suburban**<br>**%** |
| --- | --- | --- | --- | --- |
| Hispanic Non-Black* | 12.0 | 1.0 | 31.6 | 2.2 |
| Non-Hispanic Black** | 24.0 | 5.0 | 63.2 | 10.9 |
| Non-Hispanic White** | 3 | 36 | 7.9 | 78.3 |
| Committed Relationship** | 23 | 44 | 60.5 | 95.7 |
| Bachelor or above** | 6 | 28 | 15.8 | 60.9 |
| Nuliparous | 12 | 19 | 31.6 | 41.3 |
| Obese | 17 | 13 | 44.7 | 28.3 |
| Gestational Diabetes | 0 | 2 | 0.0 | 4.3 |
| Pre-eclampsia | 0 | 2 | 0.0 | 4.3 |
|  | **Average** |  | **StDev** |  |
| Age** | 28.6 | 32.3 | 5.5 | 3.0 |
| BMI | 32.0 | 28.9 | 8.2 | 6.0 |

Levels of Perceived Stress assessed by PSS-10 and the PSS-10 subconstruct Emotional Distress (“*perceived helplessness*” or “*perceived distress*”) differed by trimester and cohort, while levels of the PSS-10 subconstruct Self-Efficacy (“*perceived self-efficac*y”^35^ or “*perceived coping*”^36,37^) did not (**Table 2, Fig.1**). Overall and in the third trimester, suburban women reported higher Perceived Stress compared with urban women^32^ (overall moderate stress, PSS-10≥ 14, 56.5% versus 36.8%, p<0.01; total PSS-10 scores, 14.4 versus 10.3, p<0.001; third trimester: 59.5% versus 16.7%, p<0.01; total scores, 15.3 versus 9.1, p<0.01). There were no differences in self-reported Self-Efficacy levels between cohorts or trimesters, but White women reported higher levels of Emotional Distress in contrast to women of color, independent of the gestational trimester (9.2 versus 4.8, p<0.001).

**Table 2.** Perceived stress, Emotional Distress and Self-Efficacy levels in the two sites. While there were several statistically significant differences in total Perceived Stress and Emotional Distress between the cohorts, we didn’t identified any statistically significant differences between self-reported levels of Self-Efficacy or anxiety as measured by GAD-7. We didn’t identified any statistically significance differences in the percentage of participants that self-reported moderate/severe depression symptoms based on EPDS>=12 (suburban) or PHQ-9>9 (urban). N, number of participants; MDD, Major Depression Disorder; PHQ-9, Patient Health Questionnaire 9-items; EPDS, Edinburg Postpartum Depression Score; GAD-7, Generalized Anxiety Disorder 7-items; Gestational weeks at the T2 [24, 29] and at the T3 [33,38] visits. ^**^fdr-corrected p <0.001; ^*^fdr-corrected p <0.01.

|  | T2 and T3 combined |  |  |  | T2 |  |  |  | T3 |  |  |  |
| --- | --- | --- | --- | --- | --- | --- | --- | --- | --- | --- | --- | --- |
|  | Urban | Suburban | Urban | Suburban | Urban | Suburban | Urban | Suburban | Urban | Suburban | Urban | Suburban |
|  | N |  | % |  | N |  | % |  | N |  | % |  |
| MDD (PHQ-9 ≥ 9 or EPDS ≥ 10) | 4 | 6 | 10.5 | 13 | 2 | 4 | 8.0 | 16.0 | 3 | 6 | 7.1 | 14.2 |
| GAD-7 ≥ 9 | 4 | 6 | 10.5 | 13 | 4 | 1 | 16.0 | 4.0 | 3 | 5 | 7.1 | 11.9 |
| PSS-10 ≥ 13* | 14 | 26 | 36.8 | 56.5 | 11 | 7 | 44.0 | 28.0 | 7 | 25 | 16.7 | 59.2 |
| MDD & GAD7 | 3 | 4 | 7.9 | 8.7 | 1 | 1 | 4.0 | 4.0 | 3 | 3 | 7.1 | 7.1 |
| MDD & PSS | 3 | 6 | 7.9 | 13 | 1 | 2 | 4.0 | 8.0 | 2 | 6 | 4.7 | 14.2 |
| MDD & GAD7 & PSS | 2 | 4 | 5.3 | 8.7 | 0 | 1 | 0.0 | 4.0 | 2 | 3 | 4.7 | 7.1 |
| GAD7 & PSS | 3 | 6 | 7.9 | 13 | 3 | 1 | 12.0 | 4.0 | 2 | 5 | 4.7 | 11.9 |
|  | Average |  | StDev |  | Average |  | StDev |  | Average |  | StDev |  |
| GAD-7 | 2.7 | 3.8 | 3.7 | 3.9 | 2.7 | 2.9 | 3.8 | 3.3 | 2.6 | 4.4 | 3.7 | 4.2 |
| PSS-10* | 10.3 | 14.4 | 7.6 | 6.5 | 11.5 | 12.8 | 7.3 | 6.6 | 9.1 | 15.3 | 7.9 | 6.2 |
| Emotional Distress** | 4.8 | 9.2 | 5 | 4.2 | 5.4 | 8.2 | 5.2 | 4.4 | 4.1 | 9.7 | 4.8 | 4.0 |
| Self-Efficacy | 10.5 | 10.8 | 4.7 | 2.8 | 10.0 | 11.4 | 5.0 | 2.7 | 11.0 | 10.4 | 4.4 | 2.8 |
| PSS-10 (reduced)** | 6.6 | 10.2 | 5.4 | 4.5 | 7.5 | 9.1 | 5.3 | 4.7 | 5.7 | 10.8 | 5.5 | 4.3 |
| Emotional Distress (reduced)** | 4.1 | 7.8 | 4.3 | 3.4 | 4.7 | 7.1 | 4.5 | 3.6 | 3.5 | 8.2 | 4.1 | 3.3 |
| Self-Efficacy (reduced) | 5.4 | 5.6 | 2.5 | 1.5 | 5.1 | 6.0 | 2.7 | 1.5 | 5.8 | 5.3 | 2.3 | 1.5 |

**Figure 1.**
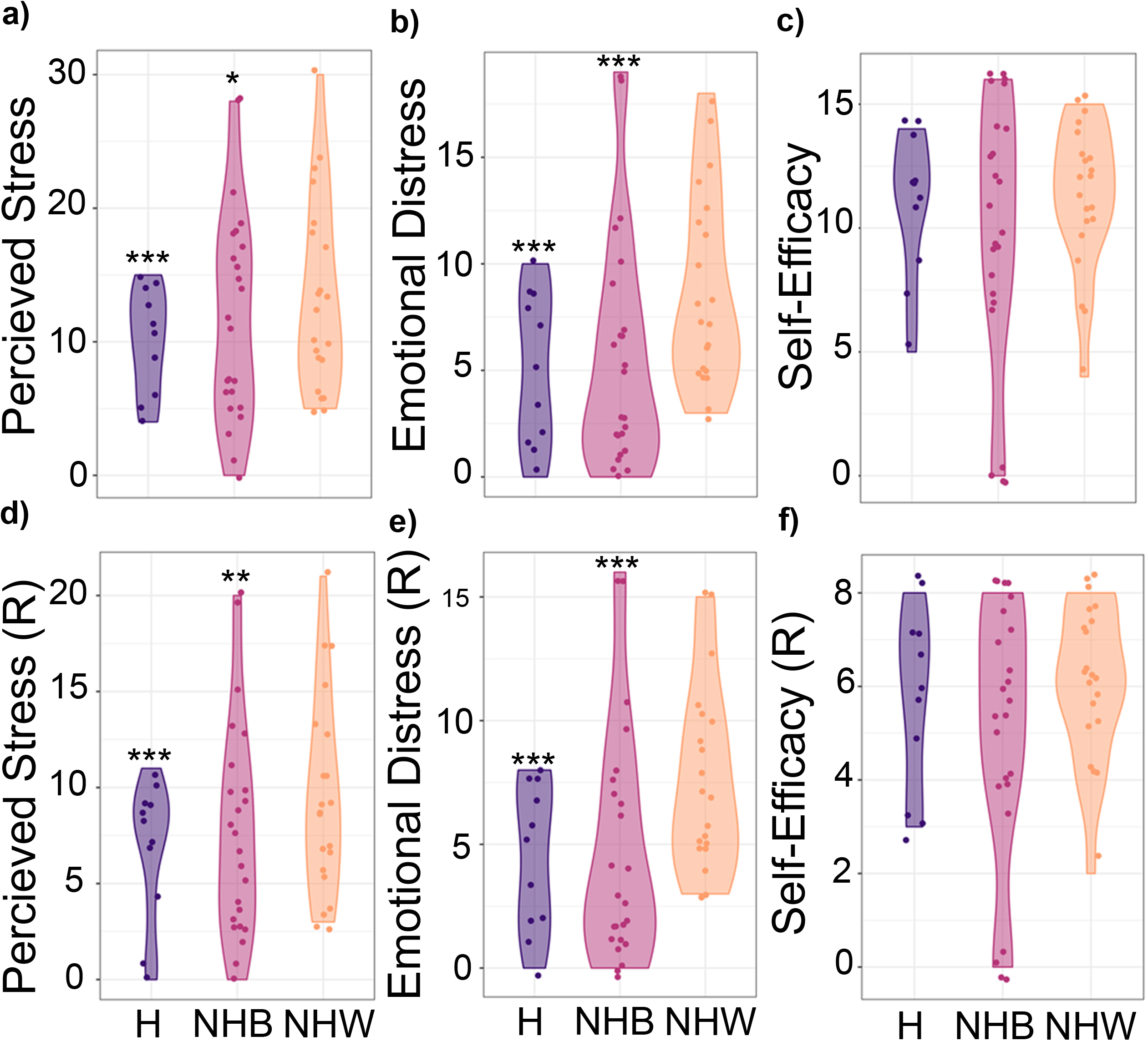
Total Perceived Stress (PSS-10), Emotional Distress and Self-Efficacy scores, including the reduced form of PSS-10 per race and ethnicity. While the levels of Total Perceived Stress and Emotional Distress (a, b) as well as their reduced forms (d, e) were higher in non-Hispanic White individuals, we observed no statically significant differences in Self-Efficacy (c) nor in the reduced Self-Efficacy values (f). R, reduced; H, Hispanic/Latina; NHB, Non-Hispanic Black; NWH, Non-Hispanic White; *p<0.05; **p<0.01; ***p<0.001

### Confirmatory Factor Analysis in the urban and suburban cohorts

To increase rigor, we explored whether measurements of self-reported Emotional Distress and Self-Efficacy were consistent between cohorts. While PSS-10 could be explained by a two factor model as previously reported^35^ (p=0.86; comparative fit index (CFI) = 0.99; Tucker-Lewis Index (TLI)= 0.99; root mean square error of approximation (RMSE)<0.001, **Fig. S1a, Table S1**), the confirmatory factor analysis indicated that the loading of the PSS-10 items in each factors, Emotional Distress and Self-Efficacy (eqs. 1-2), were very distinct between the two cohorts (weak invariance, p=0.002, **Table 3, Fig. S1b, c**). In addition, the balance between Emotional Distress and Self-Efficacy was inconsistent between the two cohorts; for a given Self-Efficacy score, the urban participants reported lower levels of Emotional Distress compared with the suburban participants (**Fig. S2**).

**Table 3.** Confirmatory Factor Analysis results. The confirmatory factor analysis established that the two factor model of Perceived Stress is not invariant and depends on the cohort. Df: degrees of freedom; Chisq, chi-square values; Chisq.diff, differences between the chi-square values for the two models; Df. diff, difference between the degrees of freedom; configural model, latent model with two factors without any constrain; weak model, constraining the loads of each item in each factor to be equal between groups; strong, constraining the loads and the intercepts for each item in each factor to be equal between groups; strict model, constraining the loads, the intercepts and residuals of the latent model for each item in each factor to be equal between groups.

|  | **Df** | **Chisq** | **Chisq.diff** | **Df.diff** | **p-value** |
| --- | --- | --- | --- | --- | --- |
| Configural | 68 | 61.428 |  |  |  |
| Weak | 76 | 99.694 | 24.187 | 8 | 0.002 |
| Weak | 76 | 99.693 |  |  |  |
| Strong | 84 | 92.639 | -13.648 | 8 | 1.000 |
| Strong | 84 | 92.639 |  |  |  |
| Strict | 94 | 146.242 | 41.053 | 10 | <0.001 |

We explored whether a reduced version of the PSS-10 enabled reporting of the same constructs of Perceived Stress, Emotional Distress and Self-Efficacy in both populations (strict invariance). Using an iterative step approach, we removed an item at a time to arrive to a significant reduced PSS-10 with seven total items. The strict two-factor model of the reduced PSS-10 was significant (p=0.73; CFI= 0.93; TLI=0.93; RMSE <0.001; **Fig. S2a, Table S2**) and was strictly invariant between the urban and suburban cohort (strict invariance p=0.14; **Table 4**). The items removed from PSS-10 to render an invariance model were *item f* from the Emotional Distress factor (‘*In the last month, how often have you found that you could not cope with all the things that you had to do?*’) and *items e* and *h* from the Self-Efficacy factor (‘*In the last month, how often have you felt that things were going your way?*’; ‘*In the last month, how often have you felt that you were on top of things?*’).

**Table 4.** Confirmatory Factor Analysis results of reduced PSS-10 questionnaire. The confirmatory factor analysis established that the reduced two factor model of Perceived Stress is invariant between cohorts. Df: degrees of freedom; Chisq, chi-square values; Chisq.diff, differences between the chi-square values for the two models; Df. diff, difference between the degrees of freedom; configural model, latent model with two factors without any constrain; weak model, constraining the loads of each item in each factor to be equal between groups; strong, constraining the loads and the intercepts for each item in each factor to be equal between groups; strict model, constraining the loads, the intercepts and residuals of the latent model for each item in each factor to be equal between groups.

|  | **Df** | **Chisq** | **Chisq.diff** | **Df.diff** | **p-value** |
| --- | --- | --- | --- | --- | --- |
| Configural | 26 | 18.381 |  |  |  |
| Weak | 31 | 23.177 | 4.008 | 5 | 0.548 |
| Weak | 31 | 23.177 |  |  |  |
| Strong | 36 | 27.423 | 8.898 | 5 | 0.113 |
| Strong | 36 | 27.423 |  |  |  |
| Strict | 43 | 36.934 | 10.891 | 7 | 0.143 |

Levels of the reduced Perceived Stress and the reduced Emotional Distress differed by trimesters and location (**Table 2**). The suburban women reported more Perceived Stress levels than urban women in the third trimester (10.8 vs 5.7, p<0.001). For the reduced Emotional Distress score, suburban participants reported higher levels overall (7.8 versus 4.1, p<0.001), as well as in each trimester (7.2 versus 4.2; 8.2 versus 3.5, respectively, p<0.001). We didn’t observe any differences in reduced Self-Efficacy between the cohorts or trimesters.

### Perceived Stress, Emotional Distress and Self-Efficacy are associated with the gut microbiota composition

While we didn’t observe any significant differences in microbial community structures (alpha-and beta-diversity and volatility) and reported levels of Perceived Stress, Emotional Distress and Self-Efficacy levels (**Fig. S4-S7)**, the proportions of 10 amplicon sequence variances (ASVs) were significantly associated with Perceived Stress, Emotional Distress and Self-Efficacy levels and also with the Ratio between Emotional Distress and Self-Efficacy independently of the recruitment location (**Fig. 2**). *Bacteroides uniformis* and *Terrisporobacter mayombei* were negatively associated with Self-Efficacy while positively associated with Perceived Stress, and *Prevotella timonensis* was negatively linked with both Perceived Stress and Emotional Distress. No ASVs were statistical associated simultaneously with both Emotional Distress and Self-Efficacy. An unspecified ASV mapped to the genus *Clostridium sensu stricto-1* was negatively associated with Self-Efficacy scores; and an unspecified ASV mapped to the genus *Faecalitalea* was negatively associated with Emotional Distress.

**Figure 2.**
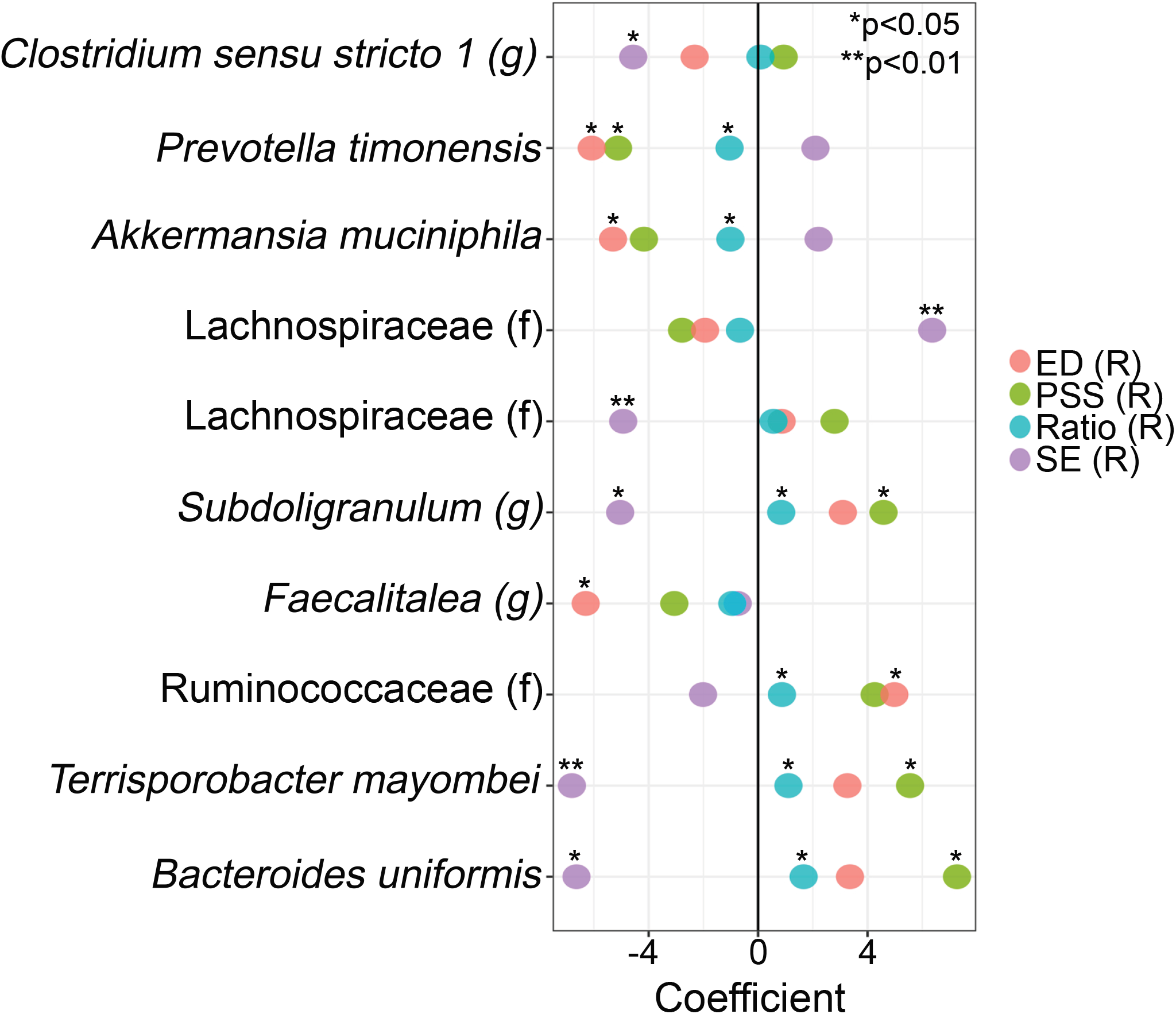
Associations between ASV with reduced Perceived Stress, reduced Emotional Distress, reduced Self-Efficacy and reduced Ratio scores. Associations between ASV normalized counts and each factor was determined using zero-inflated Generalizes Linear Models that were adjusted by recruitment site and participant’s age. Models were adjusted for multiple comparisons. R, reduced; ED, Emotional Distress; Ratio, ratio between reduced Emotional Distress score and reduced Self-Efficacy scores; SE, Self-Efficacy; *p<0.05; **p<0.01;

### Immune factors were associated with maternal socio-demographics and gut microbiome in the second trimester but not with perceived stress

We assessed T-cell related cytokine and chemokine levels in serum in the second trimester for a subset of participants (n=28). Non-Hispanic Black women had lower serum concentrations of CX3CL1 (Fractalkine), IFN-gamma, and IL-17A and greater concentrations of serum CXCL11 (ITAC) than Hispanic/Latinas and White women (**Fig. 3a-d**). While White participants, mostly from the suburban cohort (3 out 11), clustered together based on their cytokine and chemokine levels (cluster II, **Fig. 4e**), none of the measured cytokines or chemokines were associated with the perceived stress dimensions (p>0.05). Most cytokines and chemokines were positively correlated with each other, except CXCL11 (ITAC), which was negatively associated with several cytokines and chemokines including CX3CL1 (Fractalkine), TNFa, IL-5 and CCL3 (MIP-1α). The cytokines and chemokines pairs that were highly associated with each other, included IL-1b and IL-2, IL-23 and IL-21 and IL-17a and IFN-gamma (**Fig. 4f**, rho>0.9, fdr-corrected p<0.001). Similarly, hierarchical clustering of the participants by cytokine and chemokine profiles, corrected for BMI in the second trimester, resulted in three different groups of T-cell related chemokines and cytokines (**Fig. 3g**). Group I was mostly composed of inflammatory chemokines, i.e., CCL3 (MIP-1α), CCL4 (MIP-1β), CCL20 (MIP-3α), CX3CL1 (Fractalkine); Group II: anti-inflammatory cytokines (i.e., CXCL11 (ITAC), IL-4 and IL-10); and Group III, a cluster enriched in interleukins (e.g., IL-23, IL-6). The cluster with highest T-cell related cytokine and chemokine levels in serum was represented by individuals with low levels of Perceived Stress and Emotional Distress, and high levels of Self-Efficacy. On the other hand, the cluster with intermediate levels of total T-cell related cytokines and chemokines was enriched in participants that self-reported high levels in all the reduced scores, including Perceived Stress, Emotional Distress and Self-Efficacy, suggesting non-linear relationships between cytokines and chemokines levels during the second trimester of gestation and self-reported levels of Perceived Stress.

**Figure 3.**
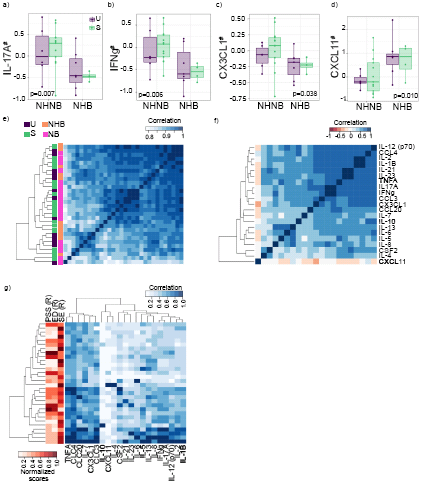
Immune system, race and reduced Perceived Stress scores during the second trimester of gestation. a-d) Several cytokines were associated with race and ethnicity— models were corrected by participants’ BMI; e) Partial Spearman correlations between cytokines and chemokine levels (log scale) corrected by participants’ BMI and level of education (as a proxy for recruitment site). We identified three main clusters, out of which, the middle cluster was greatly enriched in Non-Hispanic White participants from the suburban cohort; f) Partial Spearman correlation between cytokines and chemokines levels in serum at the second trimester correcting for participants’ BMI. While most of the cytokines and chemokines positively correlated with each other, only CCXL11 concentrations in serum were negatively correlated with the rest. g) Clustering of cytokines and chemokine (log normalized) level by participant. #Participants’ BMI was regressed out from the T-cell cytokines and chemokines log-normalized concentrations.

**Figure 4.**
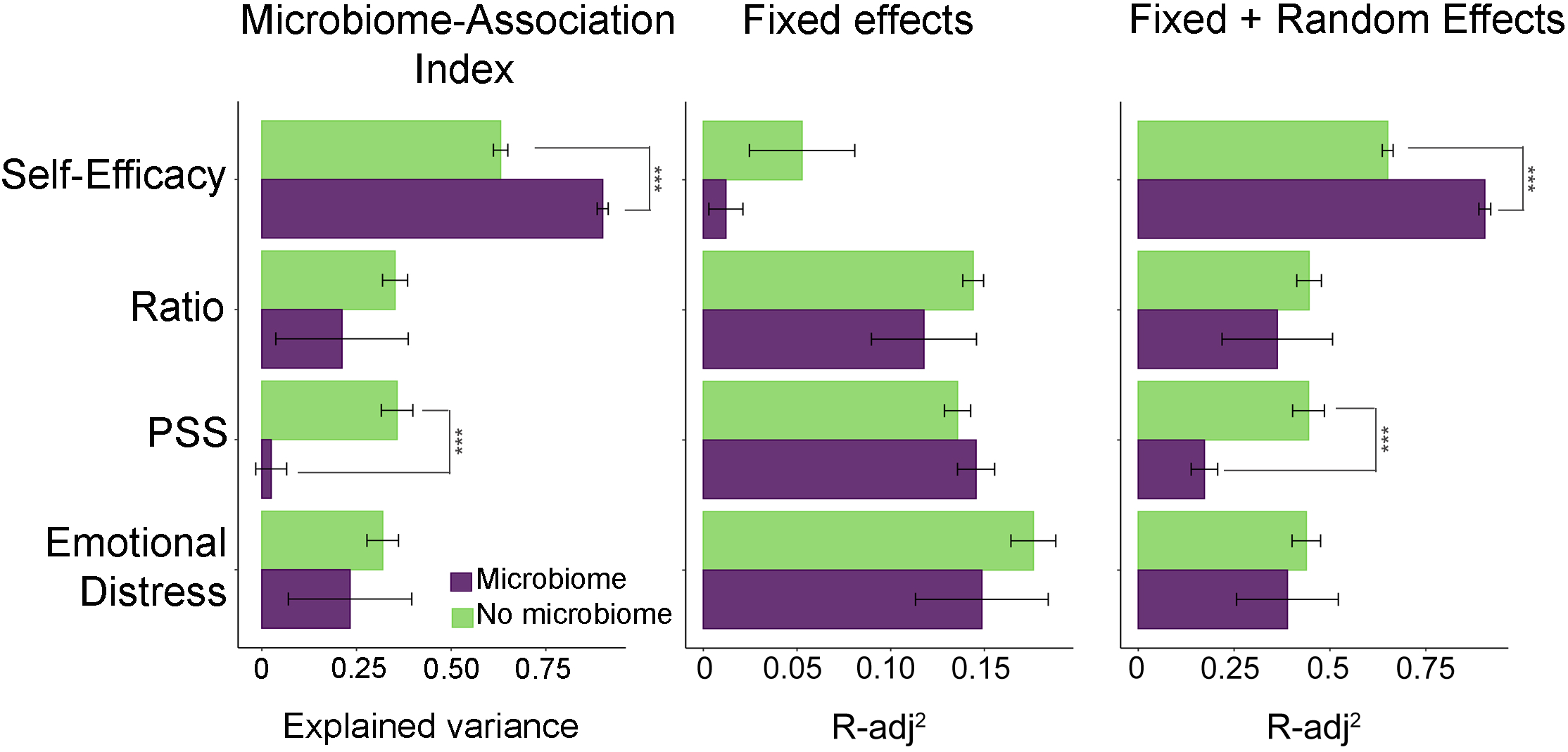
Microbiota structure predictability of self-reported Perceived Stress levels using Linear Mixed models. a) Variability of reduced Perceived Stress, reduced Emotional Distress and reduced Self-Efficacy explained by the gut microbiota composition; b) By socio-demographic values; c) Maximum explained variability by the Linear Mixed Models. Fixed factors were race/ethnicity for Emotional Distress, Perceived Stress and Ratio and nulliparity for Self-Efficacy. ***p (fdr-corrected) <0.001

When exploring the associations between the maternal circulating cytokine and chemokine levels in serum and the maternal gut microbiome in the second trimester, three immune markers, CXCL11 (ITAC), IL-5 and TNFa, were associated with seven ASVs after correcting by BMI, age and recruitment site (fdr-corrected p-value<0.005, **Table 5**). CXCL11 (ITAC) was negatively associated with *Bacteroides uniformis* and unidentified members of the genus *UCG-002* from the family Ruminococcaceae and of the family Lachnospiraceae. IL-5 was negatively associated with the genus *Methanobrevibacter* of Archaea (a methane producer) and TNFa was also negatively associated with members of the genus *UCG-002* from the family Ruminococcaceae and *Coprococcus-2*. Both IL-5 and TNFa were positively correlated with an ASV mapped to an unidentified member of the genus *Enterococcus* (**Table 5**).

**Table 5.** Significant associations between cytokines levels and ASV composition during the second trimester. Zero-inflated Generalized Linear Models were corrected by BMI, recruitment site and participant age; N=28 participants.

| **Cytokines (pg/ml)** | **Taxa** | **Coefficient** | **p-value (fdr-adj)** |
| --- | --- | --- | --- |
| CXCL11 | *f_Lachnospiraceae* | -0.02 | 5.E-03 |
| CXCL11 | *Bacteroides_uniformis* | -0.02 | 5.E-03 |
| CXCL11 | *g_Ruminococcaceae_UCG-002* | -0.03 | 5.E-03 |
| IL5 | *g_Enterococcus* | 2.53 | 3.E-03 |
| IL5 | *g_Methanobrevibacter* | -1.67 | 4.E-03 |
| TNFA | *g_Enterococcus* | 2.36 | 1.E-03 |
| TNFA | *g_Ruminococcaceae_UCG-002* | -1.10 | 1.E-03 |
| TNFA | *g_Coprococcus_2* | -1.33 | 1.E-03 |

### Predictive capabilities of the gut microbiota to explain the variations in dimensionality scores during pregnancy

Linear Mixed Models (LMM) were employed to estimate the predictive capability of the maternal gut microbiota for the self-reported scores of the reduced Perceived Stress, reduced Emotional Distress and reduced Self-Efficacy and compared them with the predictive capabilities of LMMs that included only socio-demographic factors (**Fig. 4**). More than 80% of the observed variability (microbiome association index) in the Self-Efficacy factor could be explained by the gut microbiota (p-value<0.001), significantly increasing the prediction capabilities of the LMMs by more than 25%. We didn’t observe a significant improvement in the prediction of Emotional Distress or Percieved Stress levels.

### *B. uniformis* as a potential mediator between Perceived Stress and immune system during pregnancy

Merging co-abundance correlations between the gut microbiota, immune system, and levels of perceived stress rendered multiple networks; with one of the networks containing the majority of the significant ASVs (**Fig. 5**). Within this larger network, *B. uniformis* interconnected three modules or communities characterized by immune markers, factors of perceived stress and multiple correlated ASVs. *B. uniformis* was negatively associated with the chemokine CXCL11 (ITAC) and reduced Self-Efficacy, while positively associated with reduced Perceived Stress and the ratio between reduced Emotional Distress and reduced Self-Efficacy. Although we didn’t identify any statistically significant associations between the immune system markers and the self-reported levels of perceived stress, *B. uniformis* mediated the associations between the two systems, such that CXCL11 (ITAC) was positively associated with elevated levels of reduced Self-Efficacy and inversely correlated with reduced Perceived Stress scored (note that the opposite is true for IL-5 and TNFa). The levels of Emotional Distress were negatively interconnected with several members of the maternal gut microbiota such as *Prevotella timonensis, Akkermansia muciniphila*, and a member of the genus *Faecalitalea*; but not directly to any of the measured immune markers.

**Figure 5.**
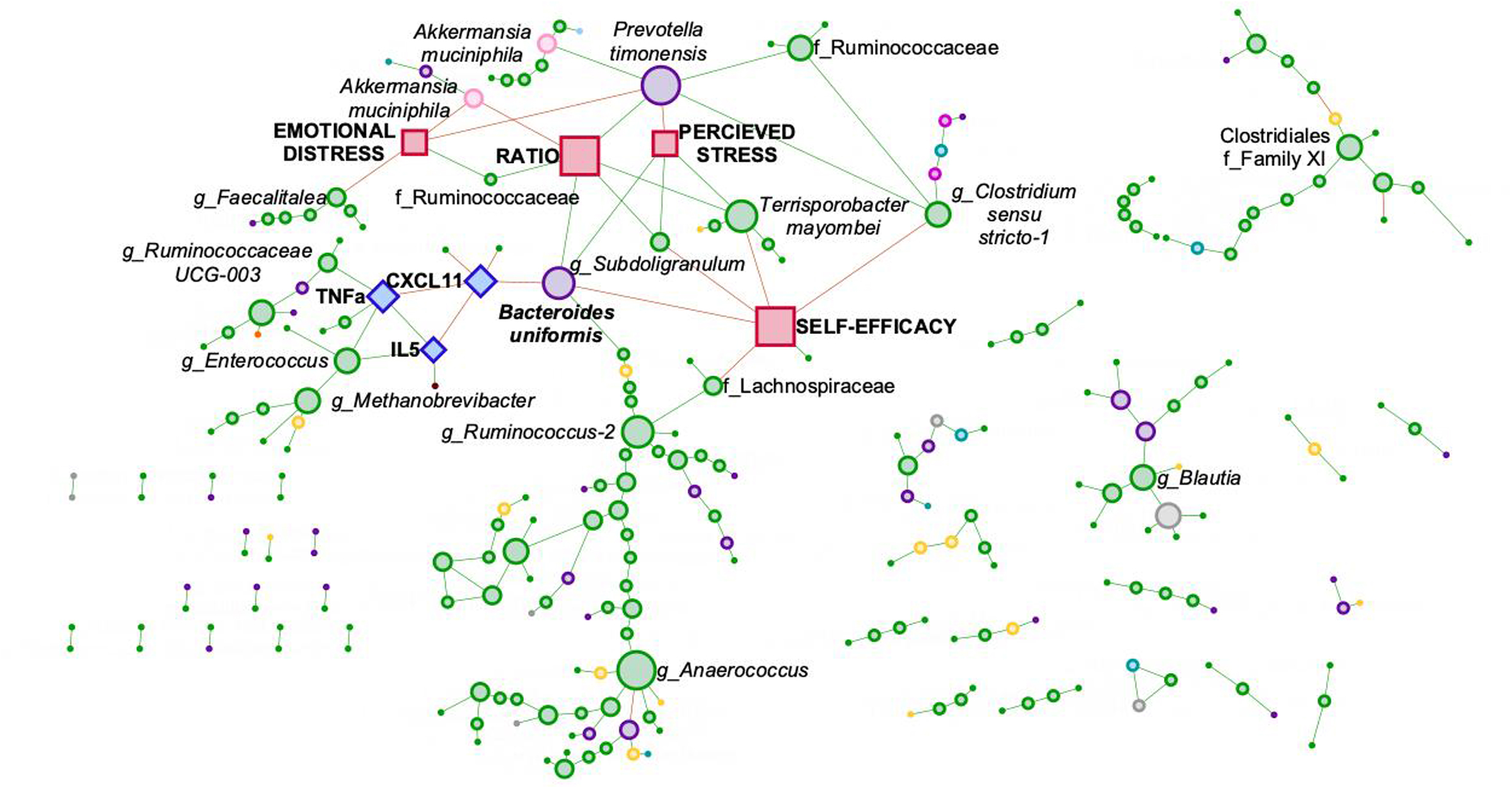
*Bacteroides uniformis* mediates the associations between the immune system, through CCXL11, and the self-reported reduced Perceived Stress assessments, through reduced Self-Efficacy and the ratio between reduced Emotional Distress and reduced Self-Efficacy. Node size is associated with its number of neighbors. Green, Firmicutes; purple, Bacteroidetes; yellow, Actinobacteria; grey, unidentified bacteria; light pink, Verrucomicrobia; blue, Protobacteria; brown, Euryarchaeota; orange, Epsilonbactereota; light blue, Tenericutes; fuchsia, Fusobacteria. Amplicon sequences variances, circles; cytokines, diamonds; stress scores, squares. Green edges, positive correlations; red, negative correlations. Associations between cytokines and chemokines with correlation coefficients were greater than 0.65 and p (fdr-corrected)<0.001 were included in the figure. Associations with **reduced** Perceived Stress, **reduced** Emotional Distress, **reduced Self-Efficaccy and reduced Ratio** were considered significance with a p (fdr-corrected)<0.05. Associations between ASV were calculated with SparCc.

## DISCUSSION

Perceived Stress is a complex construct that involves a person’s responses to the environment (Emotional Distress) and a person’s coping mechanisms (Self-Efficacy) and affects the regulation of the microbiota-gut-brain axis (MGBA). Our work sought to explore potential associations between Perceived Stress, Emotional Distress and Self-Efficacy and well-known elements of the MGBA, such as maternal inflammation and maternal gut microbiota. Our results identified links between distinct taxonomical and immunological signatures and Perceived Stress, Emotional Distress and Self-Efficacy during mid and later pregnancy.

Emotional Distress and Self-Efficacy factors measured different concepts of Perceived Stress in the urban and suburban cohorts. Specifically, the three questions that varied between our two cohorts relate to the impact of external forces as stressors (PSS-10, item f) and the individual’s sense of their ability to manage these stressors (PSS-10, items e and h). Similar results have been reported by Santiago, et. al.. The authors found that a two-factor structure of PSS-10, Emotional Distress and Self-Efficacy, was not consistent in a group of Aboriginal Australians^37^ who might face external stressors and inequalities related to colonization and historical racism that cannot be overcome by adequate coping abilities. This may suggest that differences in individual’s environments, differing views of what is a stressor, and also dissimilarities in coping and also highlights the importance of examining structural invariance when combining psychometric data from distinct populations. For instance, the suburban cohort self-reported greater levels of Perceived Stress and Emotional Distress in contrast to the urban cohort; despite individuals in the urban cohort experience higher levels of objective stress due to structural racism and lower socio-economic status (as measured by years of education), which are all associated with higher risk of postpartum depression and sustained postpartum anxiety.^54–56^ The data-driven Woods-Giscombe’s Superwoman schema that hypothesizes that Black women are more likely to feel an obligation to manifest strength, self-control and independence, suppress emotions, succeed despite limited resources, and help others despite chronic exposure to stressors is helpful in interpreting our results.^12^ In contrast, high levels of Emotional Distress, as those reported in the suburban cohort, have been associated with higher educational status and with postpartum anxiety.^57^

The gut microbiome holds promise to be an integrated metric that reflects the individual’s environment, including close social relationships^58^ and the balance of immune activation and suppression of the individual. The manner in which a person perceives stressors directly affects the MGBA and thus the host’s immune system and gut microbiome. Our results showed that reduced Emotional Distress levels were associated with microbial community habitants that reflect a shift towards more pathogenic and less beneficial microbes. For instance, higher abundance of *Akkermansia muciniphila* are associated with optimal gut barrier function and lower prevalence of chronic disorders;^66–71^ and higher levels of the pathogenic *Terrisporobacter mayombei* with illnesses such as diabetes.^72,73^ Moreover, the maternal gut microbiota during pregnancy can predict the levels of self-reported reduced Self-Efficacy, compared to socio-demographic characteristics alone, a clear indication that the perception of the subject’s environmental stressors in light of their coping mechanisms could regulate or be regulated by the maternal gut microbiota. Also, multiple taxa priorly associated with cardiovascular disease and type 2 diabetes were linked with specific cytokines (e.g., greater *Enterococcus* and IL-5 and TNF-a, lower *Coprococcus* and TNF-a), possibly indicating a shift to a more inflammatory state.^64,65^

The gut microbiome might also mediate the non-linear associations between self-reported Perceived stress levels and T-cell related cytokine and chemokine concentrations in serum and may explain the discrepancies reported in the literature pertaining to their assocations with self-reported levels of depression or anxiety, for instance.^59–63^ Our data indicates that the maternal gut microbiota may be a mediator between reduced Self-Efficacy levels and the immune system. *Bacteroides uniformis* connected the immune factor CXCL11 (ITAC) and Self-Efficacy; contributing to an inverse relationship of CXCL11 to the total Perceived Stress score. As CXCL11 binds to the CXCR3 receptor, which is mostly express in Th1 cells, primarily in the presence of foreign antigens,^74^ Self-Efficacy might serve to regulate the adequate balance of the pro-inflammatory and anti-inflammatory levels during pregnancy, which is essential for the protection of mother and the tolerance of the fetus, as fetus is considered as foreign by the maternal immune system. Similarly, CXCL11 levels have been reported to be associated with higher risk of severe COVID illness, gastrointestinal inflammation and autoimmune disorders.^75–77^ As *B. uniformis* connects CXCL11 to Self-Efficacy levels, it is plausible that *B. uniformis* also assists to balance the immune system in individuals with high levels of Self-Efficacy during the perinatal period. Prior studies have reported both positive and negative effects of *B. uniformis*. For instance, elevated levels of *B. uniformis* have been linked with depression in non-pregnant individuals, results that have been reproduced in animal model,^78^ but haven’t been explored during the perinatal period. On the contrary, there is evidence supporting the use of *B. uniformis* as a probiotic to treat obesity and binge eating.^79–81^*B. uniformis* metabolizes tryptophan, the precursor of serotonin.^83^ Insulin resistance, which occurs as pregnancy progresses, is characterized by lower ratio of tryptophan to kynurenine (T/K) and lower levels of T/K ratio are associated with greater inflammatory disorders such as preeclampsia and perinatal depression.^84–87^ Tryptophan is a key regulator of neural plasticity. Adequate neural plastic is vital to the required coping and cognitive adaptations necessary for a healthy sense of Self-Efficacy.^88^ Thus, it is plausible that elevated levels of *B. uniformis* are necessary to compensate the reduction of tryptophan as pregnancy progress to maintain a healthy levels of Self-Efficacy. Also, *B. uniformis* in combination with fiber intake is associated with reduced inflammation^79^ and our group found individuals with history of anxiety may have less dietary fiber intake.^82^ High levels of Emotional Distress are associated with elevated levels of anxiety and low levels of Self-Efficacy. Thus, it is plausible that lower diet intake combined with an inadequate microbiota-gut-brain axis due to elevated levels of anxiety and that lower levels of Self-Efficacy may lead to lower levels *B. uniformis*. Whether *B. uniformis* could be a helpful probiotic supporting immune balance and Self-Efficacy in pregnancy through tryptophan levels and fiber in the diet^89^ and whether the mediation and balancing effect of *B. uniformis* between CCLX11 and Self-Efficacy will need to further investigate to determine causality.

### Strengths, Limitations, and Future Directions

To better understand the contributors of Perceived Stress that could lead to negative health outcomes across a diverse groups of individuals, combined cohorts are needed, an strength of our study. Our study combined two diverse cohorts in relation to race/ethnicity and socio-economic status and assured that the construct to measure Perceived Stress, PSS-10, was consistent between the two cohorts. These findings may be unique to our cohorts or to pregnancy, so additional studies that include women of color and women from disadvantage populations (e.g., Native Americans, undocumented immigrants) and other perinatal times, including after delivery should be explored.^36^ While our results identified possible novel pathways of the microbiota-gut-brain axis during pregnancy, our results represents associations, not causation. Larger and more densely sampled prospectively longitudinal studies are needed to identify temporal causality. Future studies of the effects of negative stress would benefit from use of a measure of objective stressor exposure (e.g., hair cortisol levels),^90^ rather than perceived stress alone. This together with a more thorough phenotyping (e.g., past psychiatric history, diagnosis by mental health professionals, diet, emotional regulation activities) and additional tools that do not require self-report (e.g., heart rate variability, cortisol levels, clinician-or partner-rated behavioral symptoms) will greatly increase our understanding of consequences of the dysregulation of the microbiota-gut-brain axis during pregnancy and pregnancy outcomes. Finally, the use of 16S rRNA amplicon sequencing with T-cell cytokine and chemokine levels is a first step, but unbiased technologies with higher resolution to analyze the gut microbiota composition and function (e.g., metagenomics, metabolomics) and immune system (e.g., RNA-seq) are needed and animal and *in vitro* models will further improve understanding of mechanism. In Research Domain Criteria (RDoc) terms, Emotional Distress and Self-Efficacy are associated with arousal constructs (arousal and regulatory systems domain) and with the self-knowledge subconstruct (social processes domain), respectively.^91,92^ Self-knowledge and arousal are related with very distinct brain circuits and so measurements of brain connections will be important to tease apart those mechanism and the role in the complex and dynamic perinatal microbiota-gut-brain axis.

## CONCLUSIONS

Stress during pregnancy can have negative long-term consequences for the mother and the infant, and, based on our results, the microbiota-gut-brain axis might play a critical role in the mechanism of the pathobiology of the balance of Emotional Distress and Self-Efficacy that results in Perceived Stress in the perinatal period. Our study highlights the advantages of using tools and analytic methods that characterize the interaction of the maternal gut microbiome and the immune system, the nuanced view of perceived stress (e.g., two factor analysis and invariance testing) and the importance of cultural differences in reporting perceived stress. This approach also resulted in identification of a novel possible mediation role of *B. uniformis* between CXCL11 and Self-Efficacy, reflecting how the microbiome may improve our understanding of the immune activation and suppression balance that must occur in responding to stressors. Furthermore, the microbiome and immune factors may be peripheral indicators of how that balance is important to neural plasticity required for Self-Efficacy, reflecting circuits important to self-knowledge that can be highly impacted by environmental factors. An approach that integrates multiple sources of information, e.g., perception, immune and microbial attributes, is critical for assessing and identifying problematic stress during the critical perinatal period in order to improve our ability to identify individuals at risk and to guide intervention.

## Data Availability

Raw data have been deposited in SRA BioProject PRJNA667109. The samples employed for this study within this project are in the Table S3

## ACKNOWLEDGMENTS

We would like to acknowledge Hannah Rackers, Jamie Steed, and Shirin Ataei for their contributions in assisting with the cohort from the University of North Carolina and Wisslead-Pezley and Shannon Dowty from their contributions to develop the initial infrastructure at the University of Illinois at Chicago that this project relayed upon. BPB was funded by the Arnold O. Beckman Postdoctoral Award and K-12 BIRCWH Award (K12HD101373). MK is supported by the NIMH K23 Training Award (K23MH110660), NARSAD Young Investigator Award from the Brain and Behavior Research Foundation and the P&S Fund. TAEM is supported by the NIMH (K99R00MH109667; R01MH122446; RF1MH120843). This work has been partially funded by the NICHD R03HD095056. REDCap application is supported though the Center for Clinical and Translational Science (CCTS) UL1TR002003.

