## Supplemental Figures for "Interactions between Perceived Stress and Microbial-Host Immune Components in Pregnancy"

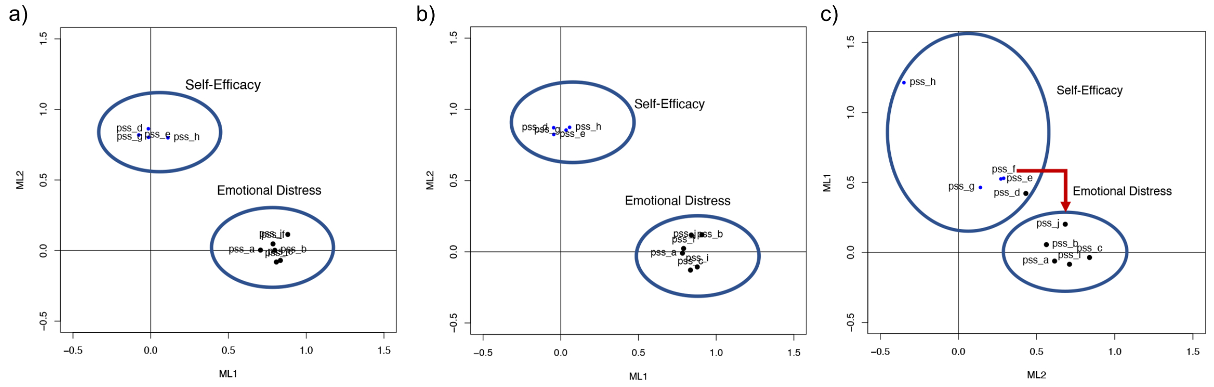


**Figure S1. Factor Analysis of PSS-10 scores. a) all participants; b) from the urban cohort; c) from the suburban cohort.** Factor analysis confirmed a PSS-10 model with two factors, Emotional Distress and Self-Efficacy, however the items associated with each factor were distinct in the urban and the suburban cohorts. For instance, item *f* from PSS-10 ("*In the last month, how often have you found that you could not cope with all the things that you had to do?*") was assigned to the Self-Efficacy factor instead of Emotional Distress in the suburban cohort. Also, for the Self-Efficacy item *h* from PSS-10 ("*In the last month, how often have you felt that you were on top of things?*"), the dimensionality reduction values in the first axis (ML1) were almost double compared with the other items of the Self-Efficacy factor in the subarban cohort (1.2 vs 0.5).

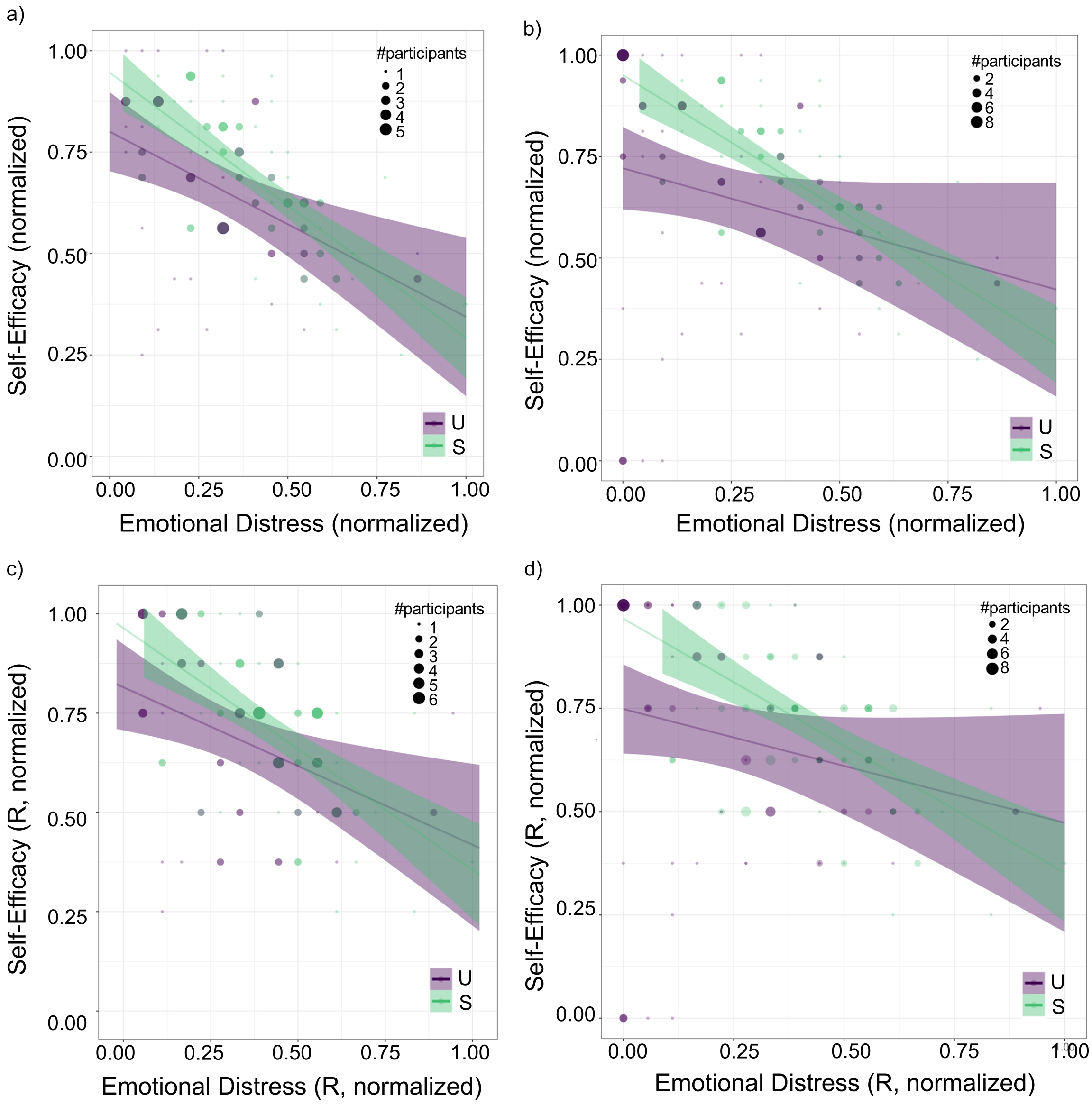


**Figure S2: Emotional Distress was inversely correlated with Self-Efficacy including all the PSS-10 items or in their reduced form for each recruitment site**. Associations between Self-Efficacy and Emotional Distress excluding outliers, participants that reported null values for self-efficacy and emotional distress (a) and including all participants (b). Associations between reduced Self-Efficacy and reduced Emotional Distress excluding outliers (c) and including all participants (d). The size of the dots is proportional to the number of participants that reported that the same levels of Self-Efficacy and Emotional Distress. The scores for each factor has been normalized between 0 and 1. R, reduced; S, Subarban; U, Urban.

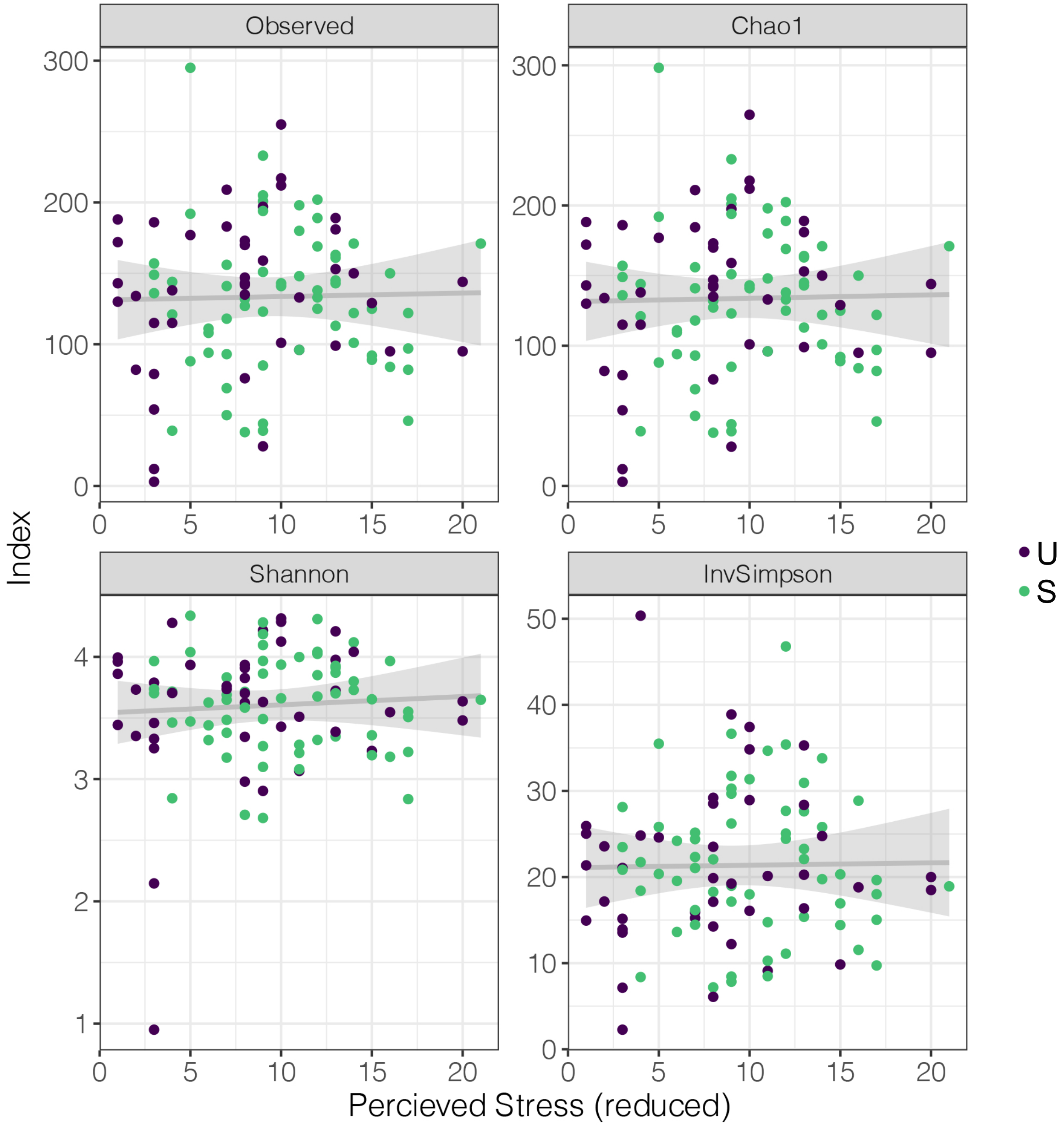


**Figure S3. The diversity and richness of the perinatal gut microbiota was not significantly associated with self-reported reduced Percieved Stress Scores.** S, suburban cohort; U, urban cohort ($p>0.05$).

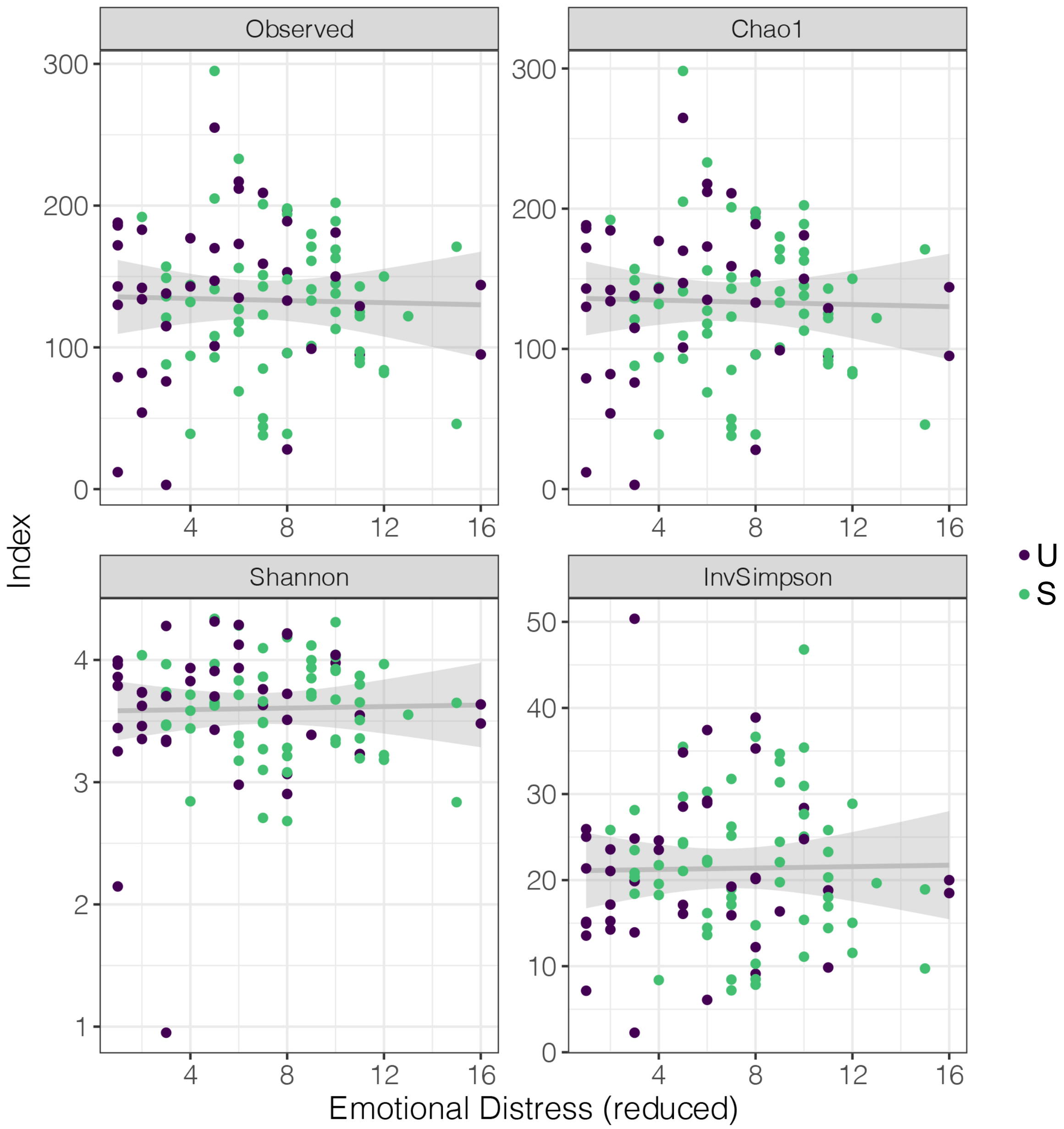


**Figure S4. The diversity and richness of the perinatal gut microbiota was not significantly associated with self-reported reduced Emotional Distress.** S, suburban cohort; U, urban cohort (p>0.05).

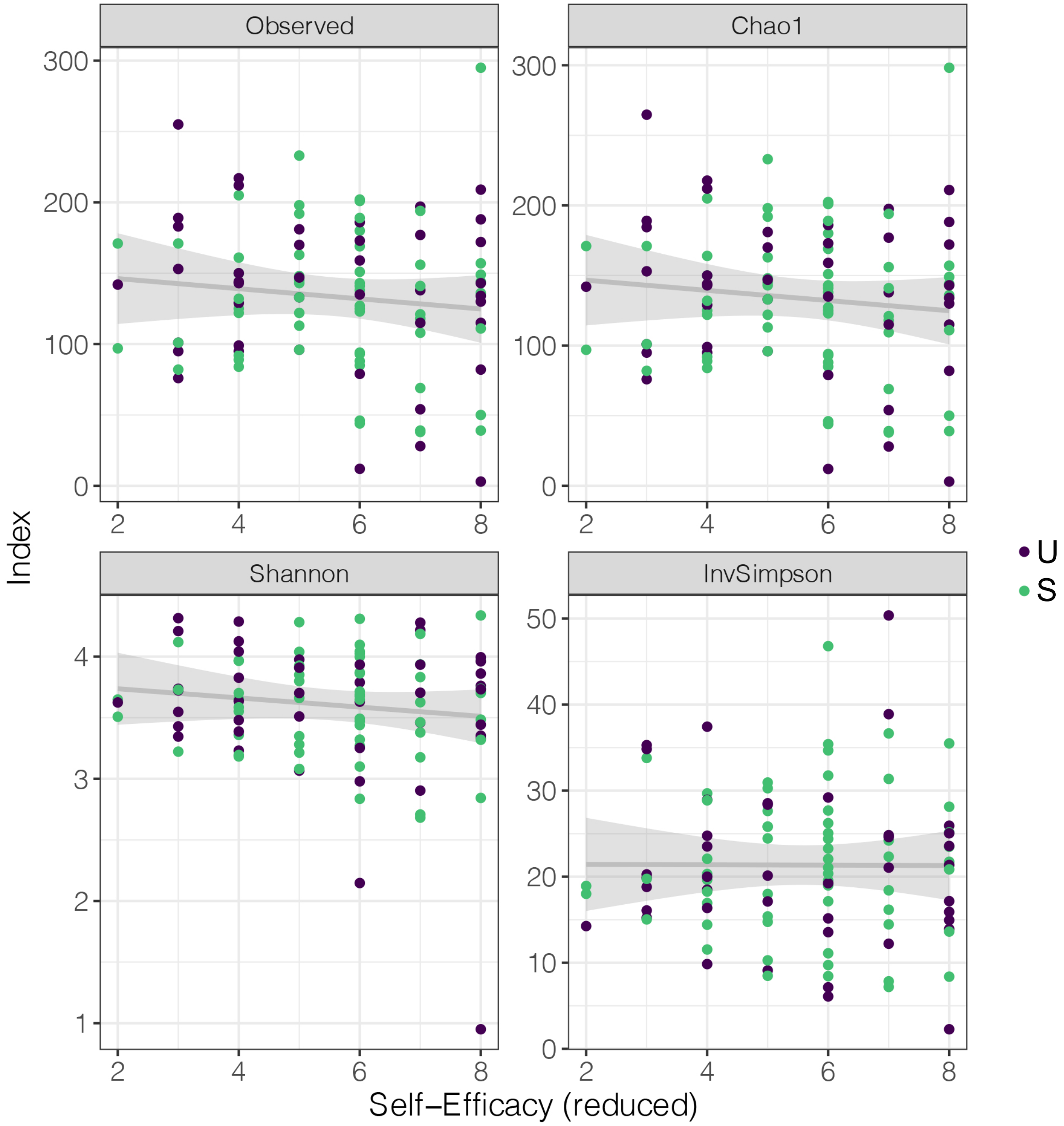


**Figure S5. The diversity and richness of the perinatal gut microbiota was not significantly associated with self-reported reduced Self-Efficacy Scores.** S, suburban cohort; U, urban cohort ($p>0.05$).

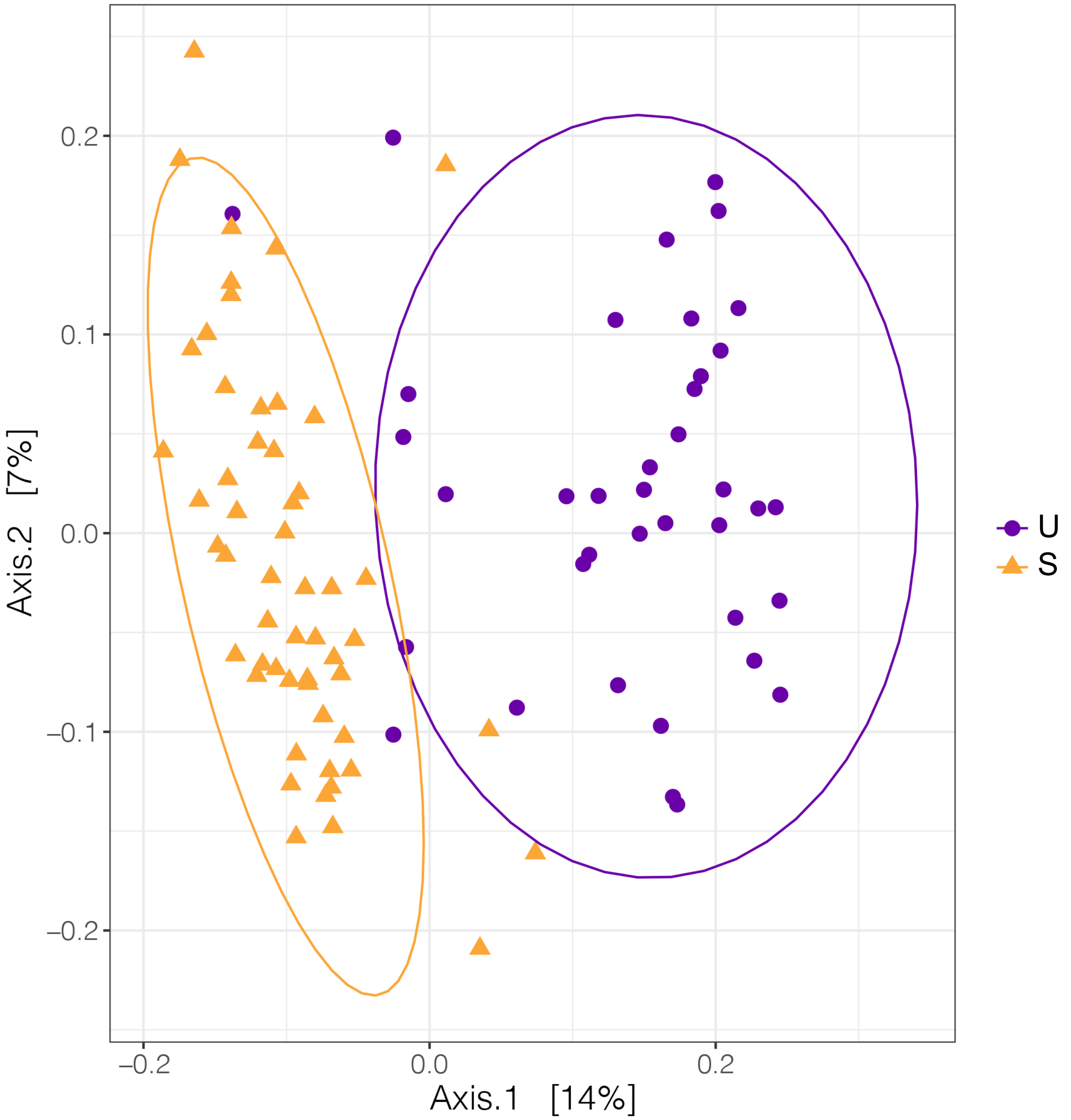


**Figure S6. The community structure of the maternal gut microbiota during pregnancy was associated with the recruitment site.** Beta-diversity was calculated using unweighted normalized UNIFRAC. S, suburban cohort; U, urban cohort (p<0.01)

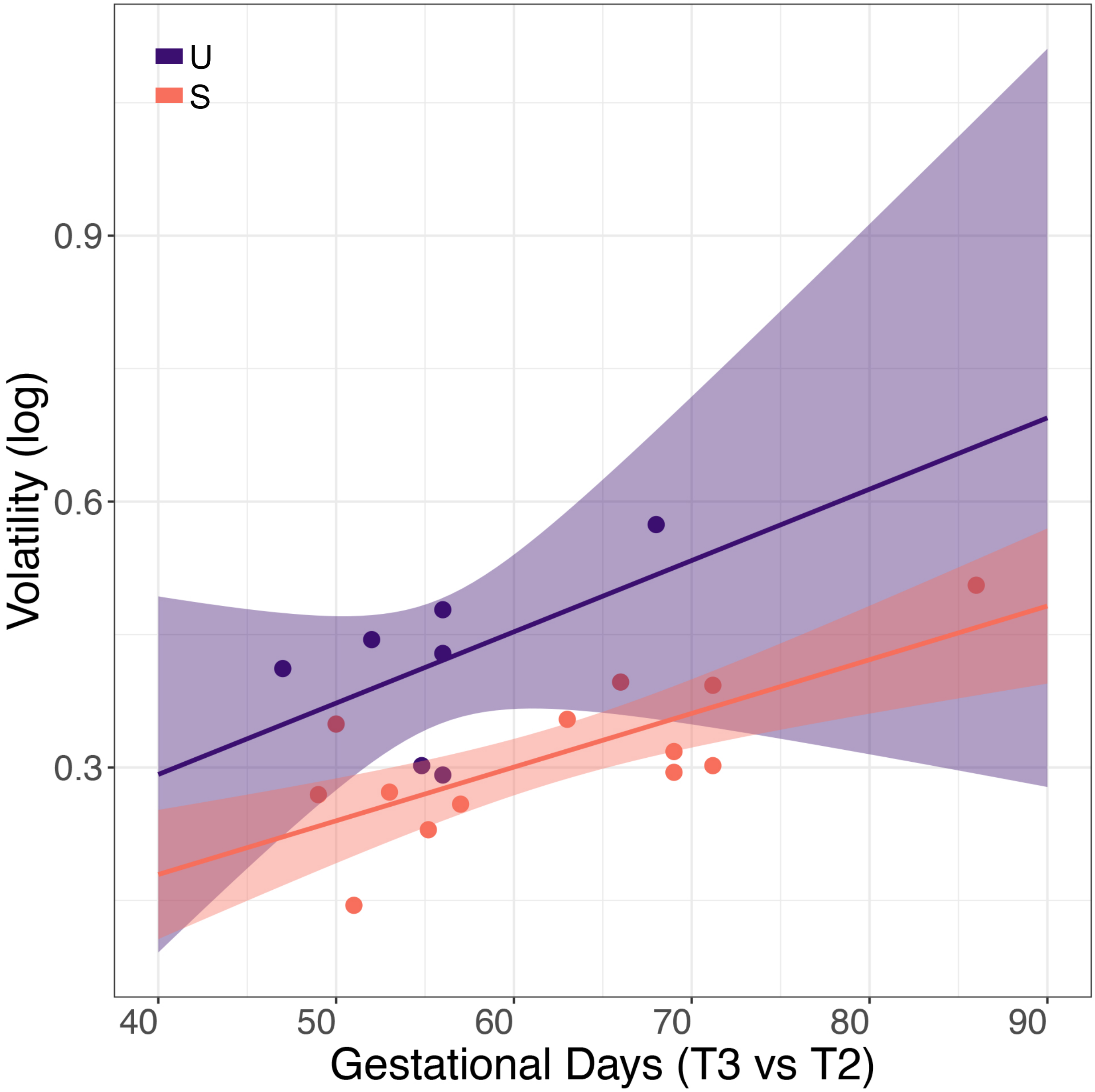


**Figure S7. Changes in volatility were associated with the days between research visits (p=0.02) and their association were dependent on the recruitment site (p=0.004).** We didn't observe any other association between volatility and any of the mental health scores of Perceived Stress. Volatility was measured as a change in unweighted normalized UNIFRAC distance. T2, second trimester; T3, third trimester; S, suburban; U, urban.
