## Supplementary material for "Interactions between Perceived Stress and Microbial-Host Immune Components in Pregnancy": Suplemental Methods

**SUPPLEMENTAL METHODS**

**Confirmatory Factor Analysis (CFA):** To compare the two PSS-10 subconstructs, Emotional Distress and Self-Efficacy, in the two cohorts, we performed CFA using latent variable modeling implemented in the R package *lavaan*.^39^ Each item of PSS-10 is scored between 0 and 4. For the confirmatory factor analysis only, “*Never” and “Almost Never”* (0 and 1) were combined as well as “*Fairly Often” and Very Often*” (3 and 4). These was necessary due to the low prevalence of individuals that reported very low values (“*Never*”, 0) or very high values (“*Very often*”, 4) for the same of the PSS-10 questions in either of the cohort that prevented the model to converge. Based on prior studies that defined the two factors of PSS-10, the following model was trained:

$$Emotional Distress=\sim pss\left( 1 \right)+pss\left( 2 \right)+pss \left( 3 \right) +pss \left( 6 \right)+pss \left( 9 \right) +pss \left( 10 \right) eq.1$$

$Self-Efficacy=\sim pss\left( 4 \right)+pss\left( 5 \right)+pss \left( 7 \right) +pss \left( 8 \right) eq.2$

$Emotional Distress=\sim Self-Efficacy eq.3$

using θ parameterization and weighted least squares means and variance adjusted (WLSMV) for parameter estimation. We explored the fitting of the two-latent variable Perceived Stress model by model configural, metric (loading), scalar steps (intercept) and residual invariances.  Models with comparative fit index (CFI) and Tucker-Lewis Index (TLI) > 0.95 and root mean square error of approximation (rmsea) <0.05 were deemed appropriate for fitting the data. Nested models were compared with ANOVA using a statistically significant p-value cutoff<0.05.

**Reduced PSS-10**: To identify the core PSS-10 items that support structural invariance between the two cohort sites (urban and suburban), we employed an iterative process in which one item was removed at a time until the model metric, scalar and residual invariability was maintained between the sites. We employed the score test or Lagrange Multiplier test, which releases one fixed or constrained parameter in the model at a time. The PSS-10 item with the lowest p-value was removed from the model until the ANOVA comparisons between the nested models was no longer statistically significant (p>0.05). Analyses were performed in R using the “*lavaan*” package.^39^

**Linear Mixed Models:** Following Rothschild and colleagues, we employed linear mixed models (LMM) to identify the predictability power of the gut microbiome data (Microbiome Associated Index) of the different dimensions identified with factor analysis.^51^ We excluded cytokine and chemokine measurements as their concentrations were only assessed in a subset of participants in their second trimester visit. We combined all the samples independently of gestational trimester, as the gestational weeks were not associated with any of the identified dimensions. Normalized ASV counts were summarized at the genus levels to construct the taxa kinship matrix (*KM*), in which the taxa kinship between participants *i* and *j* is defined as:

$${KM}_{i,j}=\sum_{k}^{n} \frac{g_{i}^{k}g_{j}^{k}}{n} (eq.4)$$

where *g_i_* and *g_j_* are the presence (1) or absence (0) of each CSS normalized genus in individuals *i* and *j* for all the genus identified in the samples and *n* is the total number of genera (*k*=[1,*n*]). Genera were considered present if their normalized counts were greater than 10. We employed the R packages *lmer* and *lme4qtl*.^52^ A step-backward approach was employed for the selection of the fixed variable in each LMM. LMM for the reduced Emotional Distress score and reduced PSS-10 scored and reduced ratio between reduced Emotional Distress and reduced Self-Efficacy were corrected by race, while for reduced Self-Efficacy score were corrected by nulliparity. To compare the effect of the addition of microbiota associated index, we developed LMMs for each of the dimensions—excluding the microbiota kinship matrix, using participants as random variables (each participant had one measurement per visit). Fixed and random variable significance was assessed with the Anova and Ranova functions from the *lmerTest* R package;^53^ prediction coefficient, R^2^, with the *MuMIn* R package; and the significance of the random effects with the exactRTLS function from the *RLRsim* package.^54^ Models were validated using a 10 times cross-validation approach.
